# Comorbidity accounts for severe COVID-19 risk, but how do we measure it? Retrospective assessment of the performance of three measures of comorbidity using 4,607 hospitalizations

**DOI:** 10.1101/2021.07.02.21259898

**Authors:** David Monterde, Gerard Carot-Sans, Miguel Cainzos-Achirica, Sònia Abilleira, Marc Coca, Emili Vela, Montse Clèries, Damià Valero, Josep Comin-Colet, Luis Garcia-Eroles, Pol Perez Sust, Miquel Arrufat, Yolanda Lejardi, Jordi Piera

**Author notes:** **Correspondence:** Jordi Piera-Jiménez, Servei Català de la Salut (CatSalut), Travessera de les Corts, 131-159 (Edifici Olímpia), 08028 Barcelona (Spain).

## Abstract

**Background:** Comorbidity burden has been identified as a relevant predictor of critical illness in patients hospitalized with coronavirus disease 2019 (COVID-19). However, comorbidity burden is often represented by a simple count of few conditions that may not fully capture patients’ complexity.

**Purpose:** To evaluate the performance of a comprehensive index of the comorbidity burden (Queralt DxS), which includes all chronic conditions present on admission, as an adjustment variable in models for predicting critical illness in hospitalized COVID-19 patients and compare it with two broadly used measures of comorbidity.

**Patients and methods:** We analyzed data from all COVID-19 hospitalizations reported in eight public hospitals of Catalonia (North-East Spain) between June 15 and December 8 2020. The primary outcome was a composite of critical illness that included the need for invasive mechanical ventilation, transfer to ICU, or in-hospital death. Predictors included age, sex, and comorbidities present on admission measured using three indices: the Charlson index, the Elixhauser index, and the Queralt DxS index for comorbidities on admission. The performance of different fitted models was compared using various indicators, including the area under the receiving operating characteristics curve (AUC).

**Results:** Our analysis included 4,607 hospitalized COVID-19 patients. Of them, 1,315 experienced critical illness. Comorbidities significantly contributed to predicting the outcome in all summary indices used. The AUC for prediction of critical illness was 0.641 (95% CI 0.624-0.660) for the Charlson index, 0.665 (0.645-0.681) for the Elixhauser index, and 0.787 (0.773-0.801) for Queralt DxS. Other metrics of model performance also showed Queralt DxS being consistently superior to the other indices.

**Conclusion:** In our analysis, the ability of comorbidity indices to predict hospital outcomes in hospitalized COVID-19 patients increased with their exhaustivity. The comprehensive Queralt DxS index may improve the accuracy of predictive models for resource allocation and clinical decision-making in the hospital setting.

## Introduction

During 2020, the rapid spread of the severe acute respiratory syndrome coronavirus 2 (SARS-CoV-2) and the severity of coronavirus disease 2019 (COVID-19) led to the collapse of many healthcare systems worldwide, particularly hospital and intensive care unit (ICU) resources. Recently approved vaccines against SARS-CoV-2 infection are expected to ease hospital resources pressure.^1^ Nevertheless, in many settings, COVID-19 continues to cause high hospital demand, which requires adequate healthcare provision planning, particularly in scenarios of a shortage of resources or overburdening of hospital resources.

Early in the pandemic, underlying comorbidities were pointed to as significant prognostic factors for the development of severe illness. Initial analyses of large series of COVID-19 patients revealed that hospitalized patients with chronic conditions like diabetes, hypertension, chronic obstructive pulmonary disease, or cardiovascular diseases were more likely to develop severe COVID-19, with no consensus regarding the contribution of each comorbidity to explaining differences in COVID-19 outcomes.^2–5^

To date, various groups have proposed multivariate models for predicting hospital outcomes in COVID-19 patients. This includes a broad range of analytical approaches that typically rely on vital signs, laboratory results, comorbidities on admission, and—less frequently—results of imaging techniques.^6^ Some of them aimed to assess patients’ risk at the time of hospital admission using measures of comorbidity burden, typically based on either unweighted counts of comorbidities or the presence of specific diagnoses from a short (i.e., up to 12) list of chronic conditions.^7–14^ However, this parsimonious approach may not fully capture patient complexity and the prognostic relevance of other conditions. Alternatively, comorbidity indices (e.g., the Charlson Comorbidity Index^15^ and the Elixhauser index^16^) combine information about several comorbidities into a single score.^17,18^ This yields more complex models with a more challenging interpretation; however, it allows adjusting for multiple factors, likely improving model accuracy.^18^

In 2020, we developed a comprehensive risk index tool for hospitalized patients, the Queralt Index, which includes a measure of pre-existing comorbidities (Queralt DxS).^19^ The Queralt DxS combines and weighs more than 2,100 relevant acute and chronic diagnostic codes. Queralt DxS has shown optimal performance for risk adjustment when measuring comorbidity burden in hospitalized patients.^19^ In this study, we assessed the performance of Queralt DxS as a comorbidity measure in models for predicting outcomes in hospitalized COVID-19 patients and compared it with other widely used comorbidity measures: the Charlson and Elixhauser indices.

## Material and methods

### Data sources

Data were retrospectively retrieved from hospital administrative databases of Catalonia, a North-East region in Spain with 7.5 million inhabitants. The dataset included admissions to the eight hospitals of the Catalan Institute of Health (ICS), which provide universal health care to nearly 70% of the Catalan population, and account for approximately 30% of all hospitalizations reported in Catalonia. ICS hospitals systematically collect and store data on diagnostics and resource utilization into a centralized database. To prevent biases associated with the unprecedented hospital overburdening experienced in Spain during the first wave of the COVID-19 outbreak,^20,21^ the primary analysis was restricted to admissions occurred from June 15 to December 8, 2020. The database was locked on March 30, 2021.

Hospital admissions due to COVID-19 were identified according to the following codes of the International Classification of Diseases, 10th Revision, Clinical Modification (ICD-10-CM) system: B97.29, B97.21, B34.2, J12.81, J12.89, and the recently added code for lab-confirmed COVID-19 U07.1. We included all hospital admissions of patients who either died in the hospital or were discharged home. Records from patients transferred from or discharged to other hospitals were excluded from the analysis to ensure the completeness of clinical data during the full index hospitalization.

All data were handled according to the General Data Protection Regulation 2016/679 on data protection and privacy for all individuals within the European Union and the local regulatory framework regarding data protection. The independent ethics committee of the Bellvitge Biomedical Research Institute (IDIBELL) approved the study protocol and waived the need for informed consent, as the data were generated as part of routine clinical care and fully de-identified for analytic purposes.

### Outcomes and Predictors

The study outcome (“critical illness” in patients hospitalized with COVID-19) was a composite that included the need for invasive mechanical ventilation, transfer to the intensive care unit (ICU), or in-hospital death. The need for invasive mechanical ventilation was identified by the presence of any of the following procedures codes recorded in the hospital database for billing purposes: 5A09357, 5A09457, 5A09557, 5A1935Z, 5A1945Z, 5A1955Z, 09HN7BZ, 09HN8BZ, 0BH13EZ, 0BH17EZ, 0BH18EZ, and 0CHY7BZ.

Potential predictors considered for the analysis included age, sex, and comorbidities present on admission. The latter were measured using three multi-comorbidity indices: the Charlson Comorbidity Index, the Elixhauser Index, and Queralt DxS. In another model, we also used the 27 comorbidities included in the Elixhauser Index, each of them separately. The Charlson and Elixhauser indices were estimated using the ICD-10 coding system proposed by Quan et al.^22^ For the Charlson index, diagnostic weights were assigned based on the original formulation by Charlson et al.^15^ For the Elixhauser Index, we used the weights proposed by Moore et al.^23^ Queralt DxS belongs to a family of three indices for predicting clinical outcomes in hospitalized patients that provides a numerical value from the weighted sum of diagnostics (primary, secondary present on admission, and complications) from a list of 2,119 diagnostic code groups.^19^ In this analysis, we used an updated version of the index (version 6.0), computed using healthcare data recorded in the routine care setting between 2018 and 2019.

### Statistics

Data on demographic and clinical variables and outcomes were described as frequency and percentage, mean and standard deviation (SD), or/and median and interquartile range (IQR, defined as the 25^th^ and 75^th^ percentiles), as appropriate. The Charlson and Elixhauser indices were computed using the Comorbidity library by Gasparini et al.;^24^ Queralt DxS was generated using R functions provided in Supplementary file 1 and used according to instructions provided in Supplementary file 2. For descriptive purposes only, patients were also stratified in four risk levels (i.e., low, moderate, high, and very high) based on the 50^th^, 80^th^, and 95^th^ percentiles of Queralt DxS, as described previously.^19^

Since the primary objective was to compare the performance of each index of comorbidity when added to a given multivariate risk prediction model, we built five logistic regression models to predict the composite outcome of critical illness: a baseline model with age and sex, three models including the baseline and each of the comorbidity measures (i.e., Charlson index, Elixhauser index, and Queralt DxS), and one including the 27 diagnoses included in the Elixhauser index separately. The performance of each model was evaluated using five statistical measures of model performance: the deviance, the Akaike information criterion (AIC),^25^ the Bayesian Information Criterion (BIC),^26^ the area under the curve of the receiving operating characteristics (AUROCC) curve, and the area under the precision-recall (AUPRC) curve.^27^ The 95% confidence intervals (CI) of the AUROCC were estimated using 1,000 bootstrap samples and confirmed with the DeLong criteria.^28^ All analyses were performed using the R statistical package (version 4.0.3).^29^

## Results

### Study Population

Between June 15 and December 8, 2020, 4,607 patients were admitted to the ICS hospitals with COVID-19. Table 1 summarizes the demographic characteristics of the study population according to the incidence of critical illness (i.e., need of invasive mechanical ventilation, transfer to ICU, or death) on admission and/or during the hospitalization. The corresponding comorbidity burden, measured using the three analyzed indices, is shown in Table 2. Patients who experienced critical illness on/after admission were significantly older and had higher scores of comorbidity burden irrespective of the index used. Men more frequently experienced critical illness.

**Table 1.**
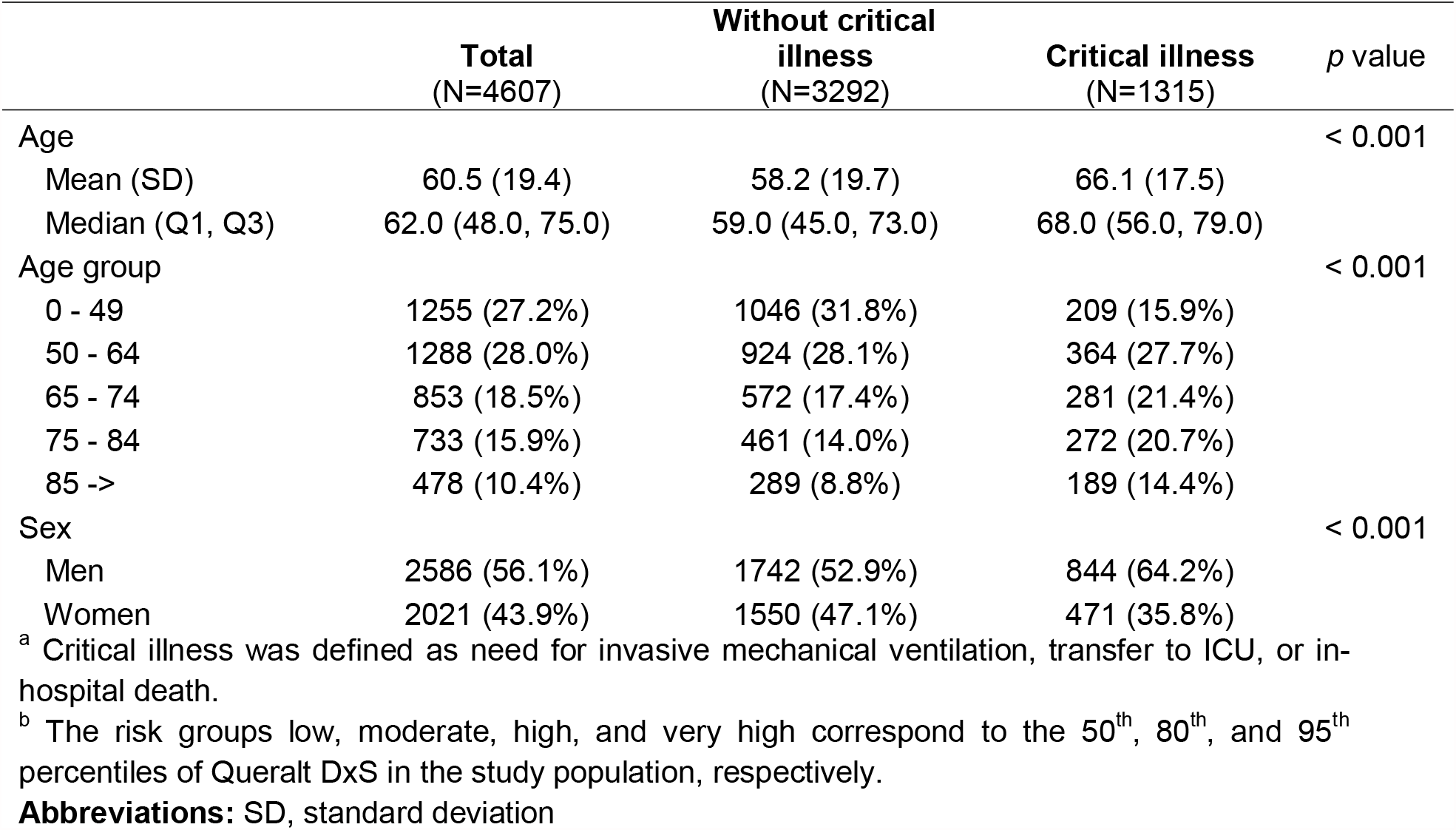
Demographic characteristics of patients included in the analysis, grouped by incidence of critical illness during the index hospitalization ^a^

**Table 2.**
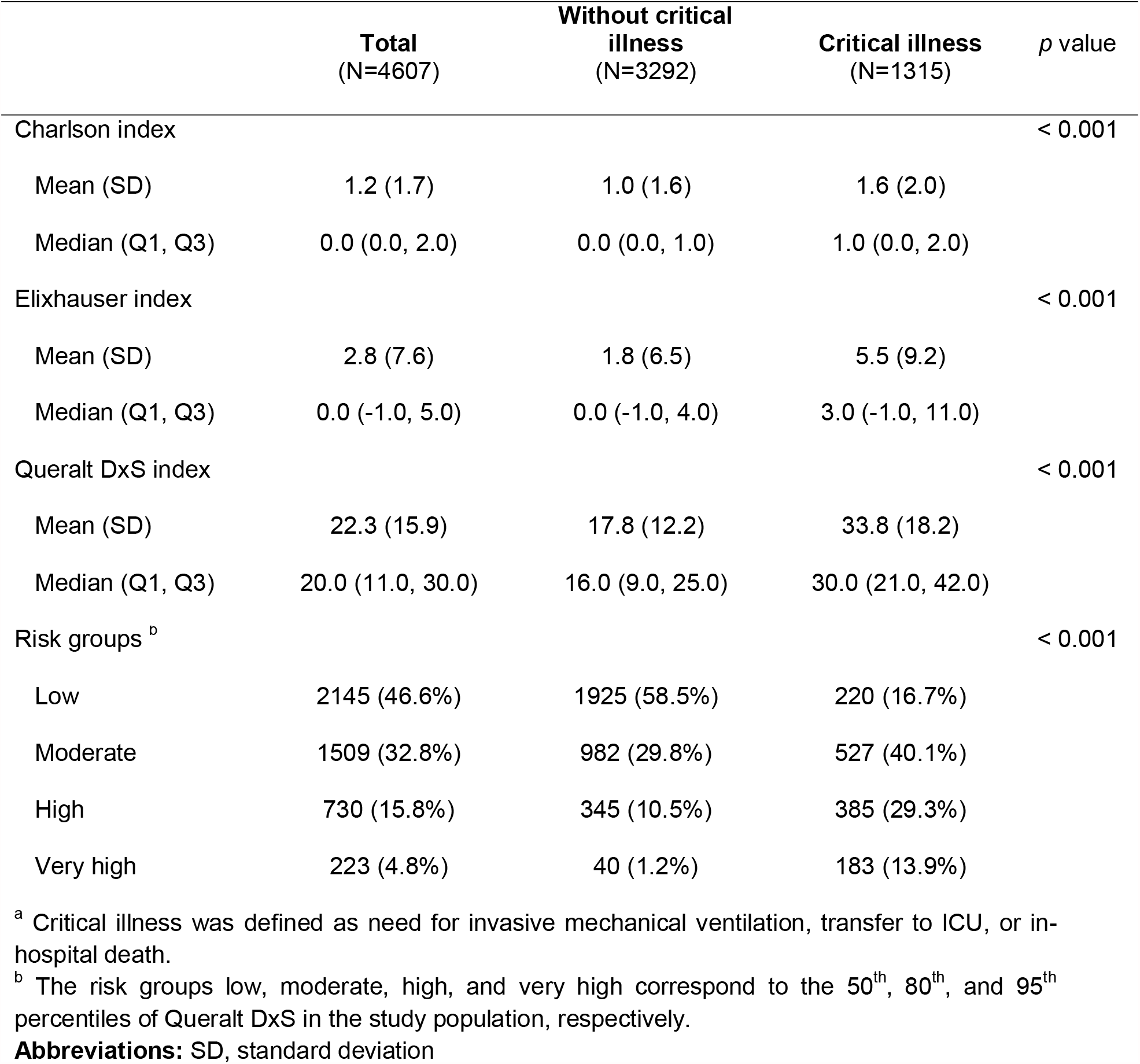
linical characteristics of patients included in the analysis, grouped by incidence of critical illness during the index hospitalization ^a^

According to the distribution of patients across the five risk levels of Queralt DxS, 2,145 (46.6%) patients were at low risk, 1,509 (32.8%) at moderate risk, 730 (15.8%) at high risk, and 223 (4.8%) at very high risk. Figure 1 shows the distribution of patients with and without critical illness, according to age, sex, and Queralt risk group.

**Figure 1.**
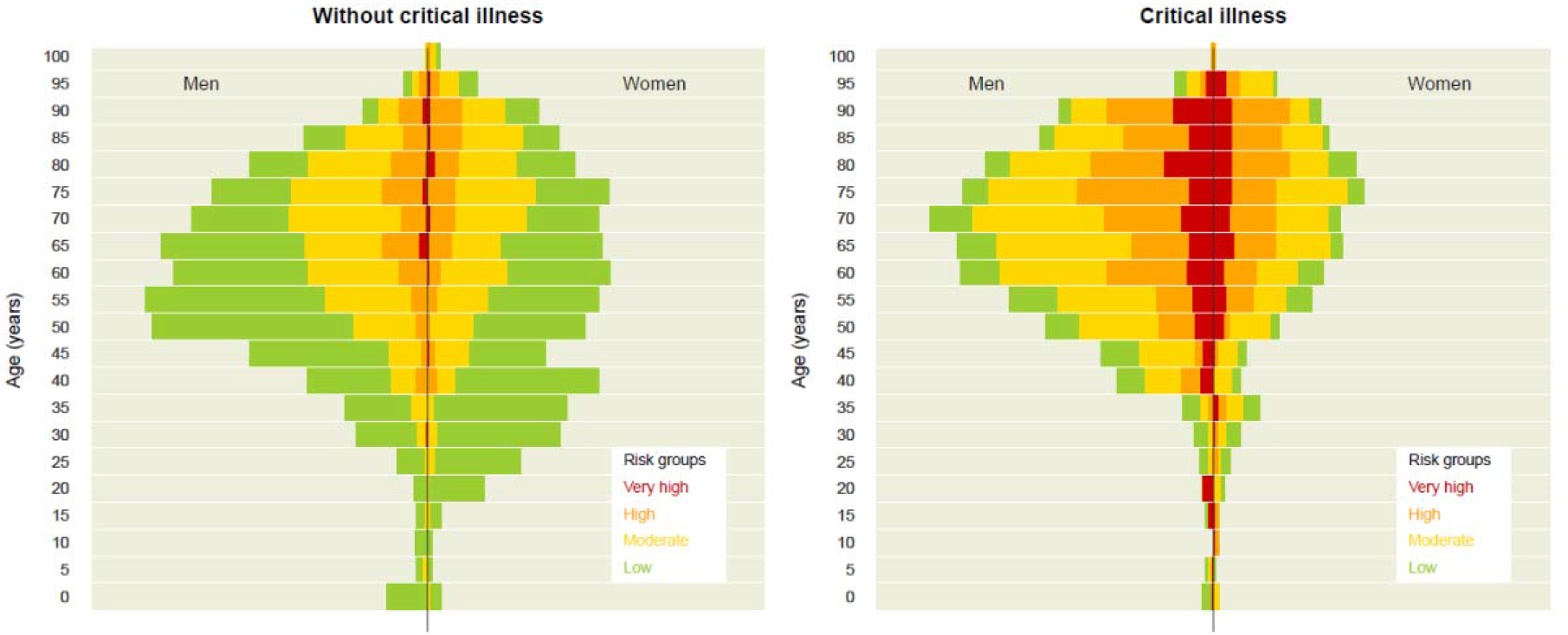
Participant distribution according to age, sex, and Queralt risk group (corresponding to the 50^th^, 80^th^, and 95^th^ percentiles of the Queralt Index).

### Performance of comorbidity indices for prediction of critical COVID-19 illness

Comorbidities significantly contributed to predicting critical illness, irrespective of the comorbidity index used in the model (Figure 2). The AUROCC was 0.641 (95% CI 0.624-0.660) for the model including the Charlson index, 0.665 (0.645-0.681) for the Elixhauser index, and 0.787 (0.773-0.801) for Queralt DxS. In the model including Queralt DxS as a comorbidity measure, age groups lost statistical significance.

**Figure 2.**
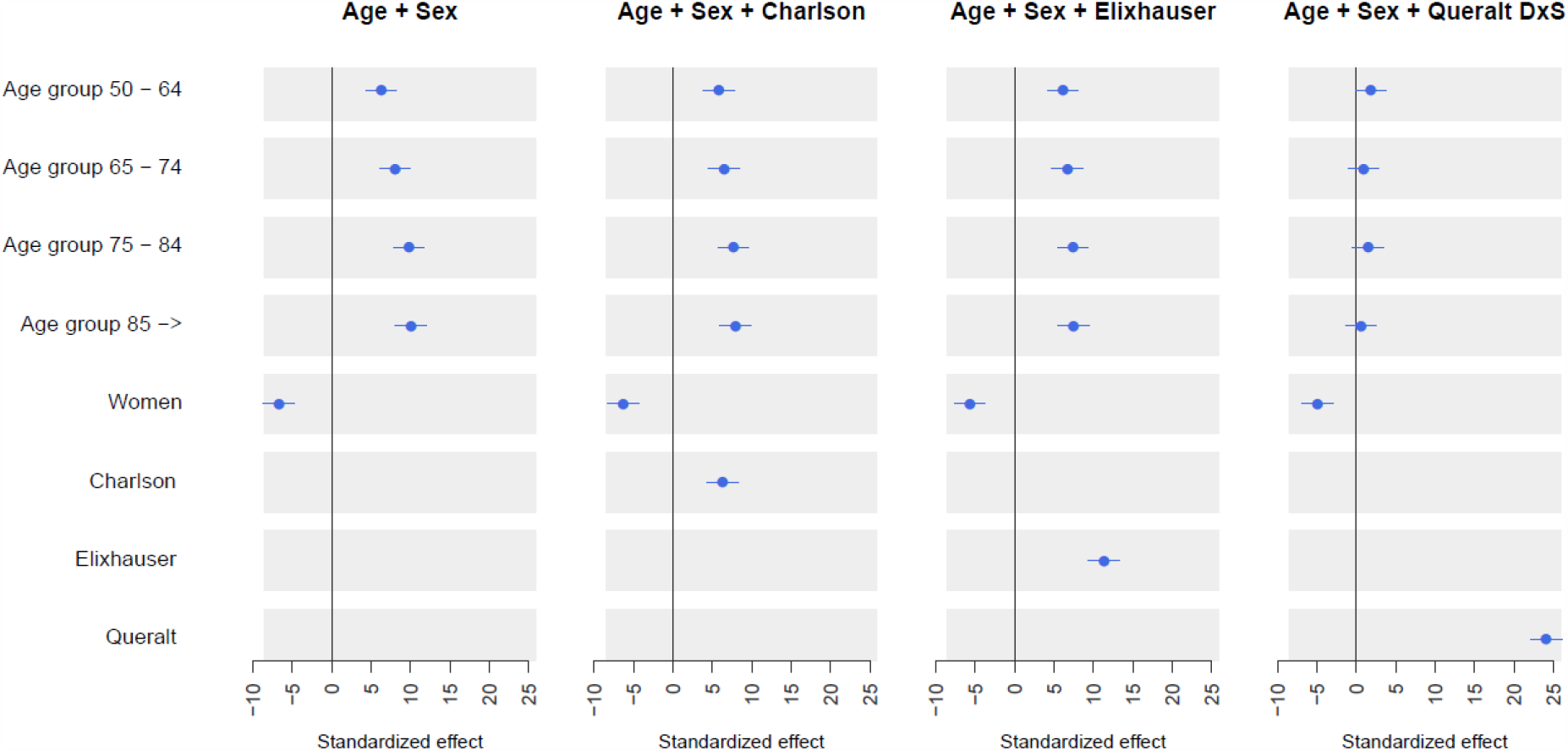
Standardized coefficients according to four logistic regression models: base (age and sex), Charlson (age, sex, and Charlson index), Elixhauser (age, sex, and Elixhauser index), Queralt (age, sex, and Queralt DxS).

The results for AUROCC were consistent with those observed using all other metrics of model performance. In all, the best performance was consistently observed for the model that included age, sex, and Queralt DxS (Figure 2). The addition of Queralt DxS to the baseline model of age and sex significantly improved the AUROCC and AUPRC of the model compared with other measures of comorbidity. Figure 3 shows the ROC and precision-recall curves of the five models explored.

**Figure 3.**
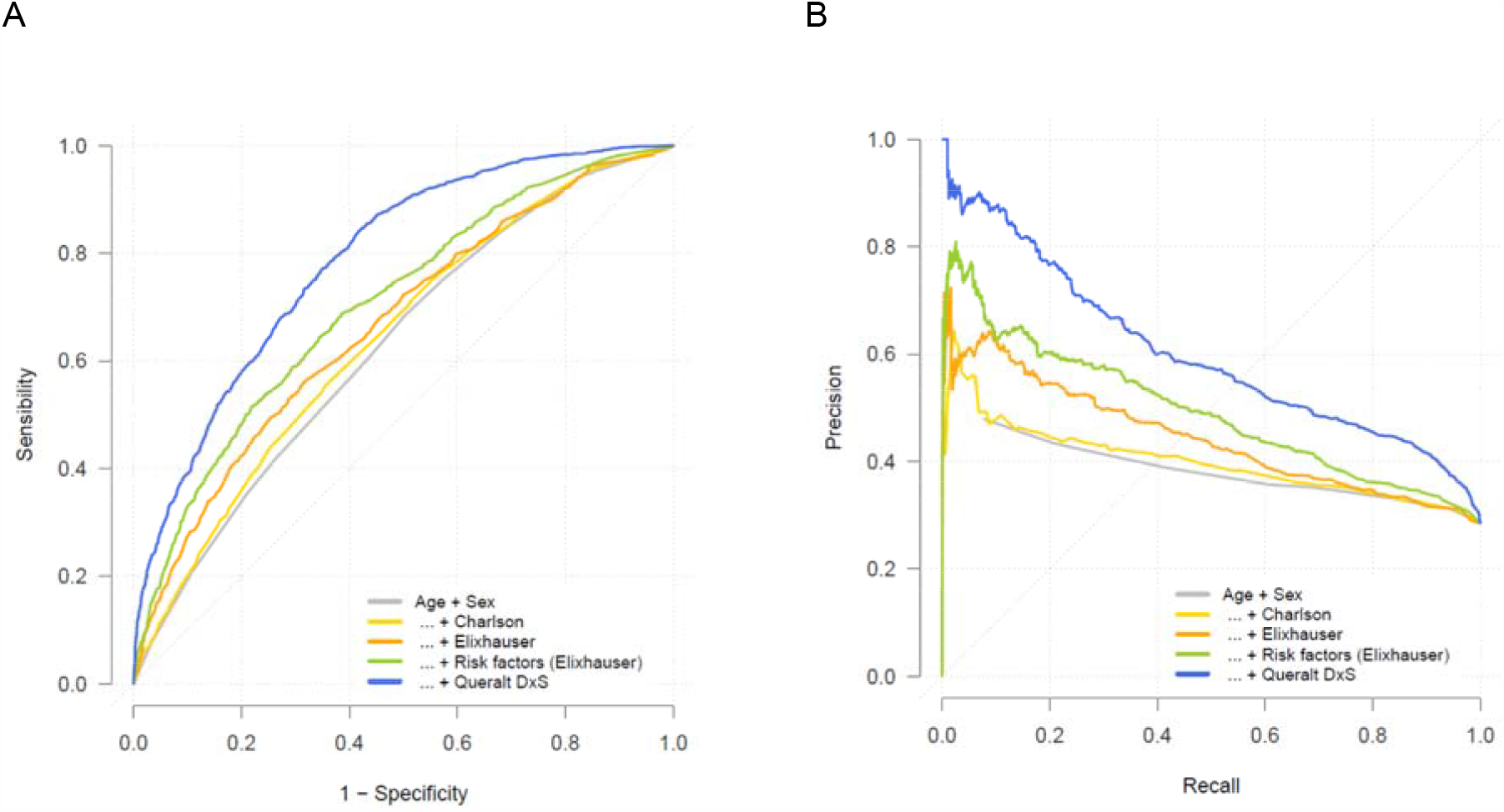
Performance of the five logistic regression models: base (age and sex), Charlson (age, sex, and Charlson index), Elixhauser (age, sex, and Elixhauser index), Queralt (age, sex, and Queralt DxS index), and the 27 diagnostic codes included in the Elixhauser index. **A:** receiving operating characteristics curve. **B:** precision-recall curve.

## Discussion

Our results using data form the healthcare databases of Catalonia confirm the relevance of comorbidities on admission as a strong predictor of outcomes in patients hospitalized with COVID-19, which is consistent with prior reports. Accordingly, the addition of any of the summary measures of comorbidity burden used in our analysis improved the performance of predictive models compared with age and sex only.

In the context of comorbidity burden being a strong predictor of outcomes in COVID-19 patients, we observed that the most comprehensive multi-comorbidity index evaluated, Queralt DxS, showed the strongest contribution to explaining critical illness. Unlike the other two indices investigated (i.e., the Charlson and Elixhauser indices), which estimate comorbidity burden from a discrete list of diagnostics, Queralt DxS considers all conditions present on admission among a list of 2,119 diagnostic groups. Hence, our findings align with the concerns highlighted by Ording and Sørensen, who warned about residual confounding potentially introduced when underestimating the comorbidity burden by using numerical indices based on rather restrictive definitions of comorbidity.^18^ Remarkably, when introducing an exhaustive summary measure of comorbidities such as Queralt DxS to the model, chronological age was no longer a significant predictor. This suggests that the risk of critical illness in older patients is driven by the accumulation of chronic conditions over time rather than by age per se.

In addition to comorbidities, various authors have identified other relevant predictors of hospital outcomes in patients with COVID-19, such as inflammatory biomarkers, need for oxygen therapy, and diagnostic images analyzed using advanced machine learning approaches.^30–35^ While these models are helpful for informing clinical decisions based on assessments performed during the hospital stay in individual patients, models that use retrospective information available from electronic health records can provide hospital outcome estimates at the time of hospital admission, thus aiding not only clinicians but also managers and policymakers in hospital resource planning. Our results suggest that all models, irrespective of whether they include data from in-hospital assessments, may benefit from a summary, highly comprehensive measure of the comorbidity burden on admission, such as Queralt DxS. We have made this index and the related code freely available online for research purposes as an open-source tool to facilitate evaluation by other groups either in its current form in other populations or potentially as part of clinical risk estimation tools combining comorbidities and clinical data on admission.

Indices that summarize a high number of variables into a single numerical index, such as Queralt DxS, also have the advantage of allowing more parsimonious models. This is particularly important for machine learning approaches, which may lose performance when increasing the number of variables included in the model. Hence, although losing sight on the individual effect of each diagnosis, summarizing the information from thousands of pre-existing diagnoses into a single index that accounts for relative weights of each and their prognostic relevance for hospital outcomes may enrich other models while minimizing the risk of overfitting.

### Study Limitations

One of the limitations of Queralt DxS is the lack validation in settings other than Catalonia, however, we hope that making the software publicly available for research purposes will facilitate external validation moving forward. Moreover, it is worth mentioning that the weights used to estimate the relative contribution of each diagnostic group to health risk were calculated using data collected during years 2018 and 2019, before the COVID-19 pandemic. Hence, although the source population of the 8 relevant hospitals has remained unchanged since then, the sample of patients admitted with COVID-19 included in this study and their clinical and sociodemographic profile do differ from those used when developing the Queralt Indices, including Queralt DxS. Therefore, the current analysis may be interpreted as a pseudo-validation of Queralt DxS in a Catalan subpopulation hospitalized with COVID-19.

Also, as often occurs in retrospective analyses, our dataset was limited to the data recorded in electronic records during routine care. Nonetheless, the universal coverage of our healthcare system, and the cross-linking of healthcare data from the primary care and specialized settings allowed us to access various sources and very comprehensive health data from our study population and consider all possible diagnostics present on admission, as well as basic demographic data such as age and sex. Conversely, we were not able to include some pre-admission variables such as blood groups, which have been associated with COVID-19 severity^36^ and might modulate the weight of comorbidities when estimating the risk of critical illness, or the SARS-CoV-2 variant, which may influence clinical outcomes of COVID-19 patients. Nevertheless, even without those pieces of information, the performance of Queralt DxS was very robust.

## Conclusion

Our findings show that the burden of pre-existing comorbidities significantly improve the prediction of risk of critical illness in patients hospitalized with COVID-19, particularly when measured exhaustively using a tool such as Queralt DxS. This comorbidity index, which is freely available for research purposes, may improve the accuracy of risk models aimed at supporting clinical decision-making and hospital resource planning in hospitalized patients, including those with COVID-19.

## Data Availability

The datasets generated and/or analysed during the current study are not publicly accessible but are
available from the corresponding author upon reasonable request.

## Abbreviations

AIC: Akaike information criterion
AUCPR: area under the precision-recall
AUCROC: area under the receiving operating characteristics
CI: confidence interval
CM: clinical modification
COVID-19: coronavirus disease 2019
ICD: international classification of diseases
ICS: Catalan institute of health
ICU: intensive care unit
IQR: interquartile range

## Ethics Approval and Informed Consent

The study protocol was approved by the independent ethics committee of the Bellvitge Biomedical Research Institute (IDIBELL), which waived the need for informed consent, as the data were generated as part of routine clinical care and fully de-identified for analytic purposes.

## Author Contributions

All authors made substantial contributions to conception and design, acquisition of data, or analysis and interpretation of data; took part in drafting the article or revising it critically for important intellectual content; gave final approval of the version to be published; and agree to be accountable for all aspects of the work.

