## Supplementary file 2 for "Comorbidity accounts for severe COVID-19 risk, but how do we measure it? Retrospective assessment of the performance of three measures of comorbidity using 4,607 hospitalizations"

### Queralt System:

#### A tool for hospital discharges risk stratification in R

User's Guide

### 1 First things first: what is Queralt System and what kind of data is expected?

#### 1.1 Queralt System: a brief introduction

Stratification tools are widely used in healthcare data world. Queralt System aims to **capture and quantify the case-mix behind hospital discharges**.

Queralt System is similar to Charlson or Elixhauser indices or Diagnosis Related Groups (DRGs): the main goal is to assign a —risk- category to a patient, based on its personal health history and the hospitalization process.

While Charlson or Elixhauser indices focuses on comorbidities and DRGs stresses on the economic dimension, Queralt System integrates both worlds and can produce **different results according to the perspective of interest**: risk, resource consumption, severity and a global score.

Also, Queralt System **can handle diagnoses and procedures**, depending on the focus of the exercise: risk adjustment or resource consumption. For the purpose of this paper, **we just provide the functions and data for diagnoses**.

The main output generated are: **groups and indices**. Groups consists of an ordinal value ranged from 1 to 5, from lowest to highest risk. **Queralt Groups** (CxD) are computed based on diagnoses groups (AD) and, thus, their aim is to stratify the risk within AD.

**Indices** are calculated as the sum of weights of subindices associated to registered ICD-10 codes, differentiating their **position and context**: principal (P), secondary (S) or present on admission (POA).

As can be seen in figure 1, subindices are also provided by the function Queralt.System.Dx(). The paper submitted uses the **IQDA_risk**, that considers POA codes and focuses on the risk of death.

The **weights** are the key component of Queralt System and are based on the data (and, hence, the characteristics) of Catalan Healthcare System users.

The algorithm considers three basic dimensions of every hospital admission: **principal diagnosis** (leading cause of the hospitalization), **secondary diagnosis** present on admission and identified thereafter and possible **hospital-associated complications**.

All in all, Queralt System considers the main elements of a hospital discharge that can be found in any administrative database, similar to DRGs, with the addition that is **open and freely distributed in R** (table 1).

**Table 1. Inputs considered by stratification algorithms**

| Inputs | Charlson | Elixhauser | DRGs | Queralt |
|---|---|---|---|---|
| Comorbidity | x | x | x | x |
| Complications | | | x | x |
| Main cause | | | x | x |
| Diagnoses | | | x | x |
| Procedures | | | x | x |
| **Outputs** | | | | |
| Categorical | | | x | x |
| Numerical | x | x | | x |

#### 1.2 The data: inputs

Queralt System for diagnoses requires a **dataframe structured in long format** with the following variables:

**Table 2. Queralt System's functions and arguments**

`Queralt.System.Dx`(Data.Dx, id = "ID", dx = "ICD", type = "Type", poa = "POA")

| Variable | Function's argument | Description |
|---|---|---|
| Data.Dx | Data.Dx | The input dataframe |
| ID | id = | Individual Id of the patient or hospital discharge |
| Diagnoses | dx = | Diagnosis codes in ICD-10 |
| Type | type = | Principal or secondary classification for the record: P or S values are expected |
| POA | poa = | Present on Admission: 0 or 1 |

It is usual that, sometimes, this kind of data is structured in wide format, that is to say, different diagnoses codes are organized through columns. When this is the case, it is possible to reshape the table via `tidyr::pivot_longer()` function.

#### 1.3 From now on

From now on it is going to be explained the main steps behind the algorithm through practical examples using the provided functions and, finally, before proceeding further, it must be made quite clear that Queralt System is for **research purposes only**.

### 2 Queralt System: diagnoses, procedures and outcomes

#### 2.1 `Queralt.System.Dx()`

As stated above, the input data must be a dataframe composed by the following columns: an Id, diagnoses in ICD-10 and without points, the type of the diagnosis (P -for principal- or S -for secondary-) and 0 or 1 if present on admission. An example data set can be found in the workspace:

```
head(Data.Dx)

    ID     ICD   Type POA
1 0872  O4292    P    1
2 0872   O669    S    1
3 0872 O99214    S    1
4 0872  E6609    S    1
5 0872  Z6830    S    0
6 0872  O9081    S    0
:   :      :     :    :
:   :      :     :    :
```

The function for the diagnoses is `Queralt.System.Dx()` and takes the following arguments (based on the example data):

```
Queralt.System.Dx(
  Data.Dx = Data.Dx,
  id = "ID",
  dx = "ICD",
  type = "Type",
  poa  = "POA"
)
```

The output consists in a new dataframe of unique identifications (ID) and joined with the main components of Queralt System; an example is shown in figure 1.

**Figure 1. Queralt System for diagnoses: outcomes**

| Result | Columns | |
|---|---|---|
| 0001 | ID | Unique Id |
| CIR019 | AD | Clinical Classifications Software Refined (CCSR) for principal diagnosis and ICD-10 chapter. |
| Heart failure | ADdesc | |
| CIR | ADM | |
| Diseases of the circulatory system | ADMdesc | |
| 4 | CxD_Global | **Queralt Groups**: an ordinal value ranged from 1 to 5, from lowest to highest risk. |
| 5 | CxD_Risk | |
| 5 | CxD_Resources | |
| 1 | CxD_Severity | |
| 45 | IQD_global | **Queralt Indices**: sum of the three main weights IQDP, IQDA, IQDC |
| 78 | IQD_risk | |
| 20 | IQD_resources | |
| 19 | IQD_seriousness | |
| 0 | IQDP_W_global | Sum of diagnoses' weights: |
| 0 | IQDP_W_risk | |
| 0 | IQDP_W_resources | |
| 0 | IQDP_W_seriousness | IQDP: Weights for Principal Diagnosis |
| 34 | IQDA_W_global | |
| 66 | **IQDA_W_risk*** | |
| 14 | IQDA_W_resources | IQDA: Weights for diagnoses present on admission |
| 11 | IQDA_W_seriousness | |
| 11 | IQDC_W_global | IQDC: Weights for diagnoses identified during hospitalization |
| 12 | IQDC_W_risk | |
| 6 | IQDC_W_resources | |
| 8 | IQDC_W_seriousness | |

*IQDA_W_risk is the input considered in the submitted paper
